# Enhancing Bayesian Kernel Machine Regression: A Dynamic Thresholding Framework to Address Variability and Skewness in High-Dimensional Environmental Health Data

**DOI:** 10.1101/2025.04.14.25325822

**Authors:** Kazi Tanvir Hasan, Gabriel Odom, Zoran Bursac, Roberto Lucchini, Boubakari Ibrahimou

## Abstract

Bayesian Kernel Machine Regression (BKMR) is widely used in environmental health research to model complex, nonlinear, and interactive relationships in high-dimensional datasets. However, using a fixed posterior inclusion probability (PIP) threshold can lead to inconsistent test size control, influenced by the coefficient of variation (CV) and sample size. This study introduces a dynamic thresholding approach that adapts to these dataset characteristics, improving the sensitivity and reliability of BKMR analyses. A four-parameter logistic regression model was developed to estimate the 95th percentile of PIP as a function of the log-transformed CV and sample size. Simulations were performed across a broad range of CV values and sample sizes to evaluate test size performance for fixed and dynamic thresholds. The dynamic threshold was validated with independent simulated datasets and applied to the 2011–2014 NHANES data. The dynamic threshold consistently maintained nominal test sizes near five percent, outperforming the fixed threshold, which exhibited substantial variability. Validation with empirical data identified cadmium, manganese, and lead as significant contributors to cognitive performance, with cadmium emerging as the most influential. The dynamic threshold approach improves the precision of variable selection in BKMR, offering a more reliable method to analyze complex exposure-response relationships in environmental health research.

## Introduction

The study of health effects from exposure to complex multipollutant mixtures is a critical area of focus in environmental health research^1–3^. Mixtures, which include air pollutants, toxic waste and persistent organic chemicals, can potentially display nonlinear and non-additive exposure-response relationships^4,5^. Such complexities, coupled with the high dimensionality of data and intricate intercorrelations among exposures, pose significant challenges to risk assessment based on traditional statistical methodologies^5–8^. Addressing these challenges requires advanced analytical frameworks capable of disentangling the complex interdependencies inherent in environmental health data^5,6,9,10^. Bayesian Kernel Machine Regression (BKMR) is a powerful tool for modelling nonlinear and interactive relationships between exposures and outcomes^5,6,11–14^. By leveraging kernel functions, such as the Gaussian kernel, BKMR provides a flexible framework for analyzing exposure-response dynamics. Its Bayesian structure further strengthens its utility in high-dimensional contexts by offering robust posterior probability estimates for predictors^5,6,12–15^. Despite these advantages, BKMR exhibits limitations, particularly in scenarios involving high variability or skewness in response data^5,6,16^. These factors can adversely impact the model’s sensitivity and reliability, necessitating enhancements to its foundational methodology.

One critical limitation of traditional BKMR is its reliance on fixed thresholds for posterior inclusion probabilities (PIP), such as the commonly used value of 0.5^14,15,17–21^. This static approach does not account for variations in sample size or the response variable’s coefficient of variation (CV), which measures relative variability in the data. As a result, test size—the probability of falsely rejecting the null hypothesis—can deviate significantly from the nominal level (e.g., 5%), leading to suboptimal statistical inferences. This shortcoming can compromise the sensitivity and reliability of BKMR analyses, particularly in high-dimensional or highly variable datasets. For instance, our previous studies^16^ have shown that small CV values can cause test size to be overly conservative, approaching 0%. In contrast, high CV values often result in anti-conservative test sizes exceeding 50%. Such inconsistencies compromise the ability of BKMR to accurately identify important predictors and highlight the inadequacy of fixed thresholds in diverse datasets^16^.

Test size plays a fundamental role in hypothesis testing, influencing the validity and reliability of statistical inferences^22–24^. Deviations from nominal test size levels may result in either conservative outcomes, characterized by reduced statistical power, or anti-conservative outcomes, where false positive rates are inflated^25^. These issues are particularly relevant in environmental health contexts, such as studies predicting cognitive decline from heavy metal concentrations in blood or urine, where data are often skewed^26–30^. Our findings suggest that relying on a fixed PIP threshold of 0.5 to infer variable importance may lead to overlooking relevant variables associated with the outcome or including irrelevant variables by mistake. The skewed data distribution introduces complexities that challenge the appropriateness of this conventional threshold, emphasizing the need for a subtle understanding of variable importance determination in skewed data scenarios^16,27,31^.

Despite these challenges, BKMR’s ability to incorporate off-diagonal covariance information, which captures interdependencies among predictors, remains a key strength^5,6^. This feature enhances statistical power and accuracy, making BKMR a valuable tool for unravelling complex exposure-response relationships^5,6,9,10^. Building on these foundational strengths, this study seeks to evaluate BKMR’s variation in test size—a fundamental property representing the probability of falsely rejecting the null hypothesis—under diverse CV and sample size conditions. Addressing these limitations is vital to enhancing BKMR’s reliability and robustness. We propose a dynamic thresholding framework to address the limitations of fixed thresholds in BKMR. By incorporating a logistic regression-based threshold function, our approach adaptively adjusts for CV and sample size variations, enhancing the method’s sensitivity and reliability.

This study makes several contributions to the advancement of health studies involving mixtures by improving the BKMR methodology:

- Performance Evaluation: We systematically evaluate BKMR’s ability to detect actual effects under varying CV and sample size conditions.
- Development of a Dynamic Thresholding Framework: Using a four-parameter logistic regression model, we introduce an adaptive thresholding approach that accounts for variability in data characteristics.
- Comparative Analysis: We compare the proposed dynamic thresholding method with the traditionally employed fixed threshold, demonstrating its superior ability to maintain nominal test size and improve sensitivity.
- Application to Environmental Health Data: We apply the enhanced BKMR framework to an environmental health dataset examining metal mixtures and cognitive function in a U.S. population.

By addressing the limitations of fixed thresholds, our proposed methodology provides a robust framework for utilizing BKMR in high-dimensional and variable-rich datasets. This research offers a significant contribution to environmental health studies and beyond, paving the way for more accurate risk assessment through reliable exposure-response analyses. The findings are anticipated to inform public health interventions and policies by improving the robustness of BKMR analyses.

## Methodology

### Overview of Bayesian Kernel Machine Regression (BKMR)

Bayesian Kernel Machine Regression (BKMR) is a sophisticated statistical approach to model complex, nonlinear, and interactive relationships between predictors and outcomes. By leveraging kernel functions, such as the Gaussian kernel, BKMR flexibly captures high-dimensional exposure-response dynamics, making it a valuable tool in environmental health studies. The BKMR model is an extension of the Kernel Machine Regression model that allows for including a mixture component^5,6^. The BKMR model is given by

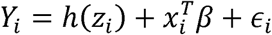

where *Y_i_* is a health endpoint, *z_i_*= (*z_i1_*,.. ., *z_iM_*)*^T^*is a vector of M exposure variables (e.g. heavy metals), X_i_ contains a set of potential confounders, and ϵ_*i*_ ^i.i.d.^_∼_ *N*(0,σ^2^)^5,6^.

Its Bayesian framework enhances reliability, offering robust posterior probability estimates and allowing for a nuanced variable importance assessment^5,6,8,12–14^. Despite its strengths, the BKMR model’s performance can vary significantly based on factors such as the response variable’s coefficient of variation (CV) and the sample size, mainly when relying on fixed thresholds like the commonly used posterior inclusion probability (PIP) threshold of 0.5^13,14,16–21^.

The primary objective of this analysis was to evaluate BKMR’s sensitivity to varying CV and sample size conditions, focusing on the test size (the probability of rejecting the null hypothesis when it is true). In our prior work16, we demonstrated that the BKMR test size varies dramatically with CV, ranging from 0% (highly conservative) for near-zero CV values to over 50% (highly anti-conservative) for CV values exceeding 10. To address this limitation, we aimed to predict the 95th percentile of PIP (*PIP_q95_*) as a function of CV and sample size, ultimately deriving dynamic threshold values to maintain a consistent nominal test size of 5%.

This analysis was conducted in three phases:

- BKMR model evaluation: Estimation of *PIP_q_*_95_ across various CV and sample size combinations using simulation training data.
- Logistic model fitting: Development of a nonlinear regression model using the Richard Curve to predict *PIP_q_*_95_ in the training data.
- Threshold comparison: Assessment of the new dynamic thresholds against the traditional fixed threshold of 0.5 using independent validation data.

### Phase 1: BKMR Model Evaluation

#### Data Inputs

Simulated datasets were created to represent a range of response variability and sample sizes:

- Coefficient of Variation (CV): 41 unique CV values ranging from 0.0235 to 15.0000. Sample Sizes: Six groups were analyzed, with sizes of 150, 300, 600, 1000, 2000, and 2934 (100% of data)^16,32–36^.

#### Model Setup

- Software/Method: The BKMR model was implemented using the kmbayes function in R. Each model was run for 10,000 Markov Chain Monte Carlo (MCMC) iterations^37^.
- Performance Metric: The 95th percentile of PIP (*PIP_q_*_95_) was computed for each combination of CV and sample size.
- Repetition: For each combination, 100 simulated datasets were analyzed to ensure robust results and reduce variability due to random factors. First, 50 randomly selected datasets were used to build the model equation (training analysis), and the remaining 50 datasets were used for model evaluation (validation analysis).
- In this analysis, we used a high-performance computing (HPC) node. The node is equipped with four Intel(R) Xeon(R) CPU E7-8890 v3 processors, each operating at 2.50GHz, providing a total of 72 CPU cores. Additionally, the node has 512 GB of RAM.

### Phase 2: Logistic Model Fitting

To understand the relationship between *PIP_q_*_95_, CV, and sample size, a four-parameter logistic regression model (Richard Curve) was employed^38^.

#### Model Specification

The four-parameter logistic equation is:

Where:

- y: the 95th percentile of the PIP values *PIP*_*q*95_
- A: Left asymptote, fixed at 0.
- K: Right asymptote, bounded above by 1.
- C: Constant.
- β_1_, β_2_: Midpoint shift parameters for CV and sample size.
- x1: Log-transformed CV *log*_2_(CV).
- x2: Log-transformed sample size *log*_10_(Sample Size).

#### Nonlinear Regression Procedure

The relationship was modeled using the nls function from the R stats package^39^, and the Levenberg-Marquardt algorithm^40–42^ was employed for optimization.

- Parameter Initialization: Initial values for C, βJ_1_, and βJ_2_ were set to 1, based on prior knowledge and typical system behavior^38,43^. Also, the value of k is fixed at 1 as the maximum of PIP.
- Residual Diagnostics: Residuals were analyzed to ensure no significant bias or deviation from randomness.
- Convergence Validation: Model assumptions and convergence were confirmed through parameter stability and goodness-of-fit measures.
- Validation Steps: Alternative initial values were tested to ensure robustness against local minima, and residual diagnostics validated the model’s reliability.

### Phase 3: Threshold Comparison

A dynamic threshold function was derived from the logistic model to adjust thresholds based on CV and sample size.

#### Dynamic Threshold Function

The dynamic thresholds were compared with the traditional fixed threshold of 0.5.

#### Dynamic Threshold Function

- Test Size Control: The ability of each threshold approach to maintain a nominal test size of 5%.
- Sensitivity: The rate of correctly identifying actual effects under varying CV and sample size conditions.

This methodology combines simulation-based BKMR evaluation, nonlinear logistic regression modelling, and comparative threshold analysis to address the limitations of fixed thresholds in high-dimensional datasets. The proposed dynamic threshold function enhances the reliability and validity of BKMR analyses, paving the way for more robust exposure-response modelling in environmental health research.

## Results

### Model Fit and Predictions

The logistic model demonstrated a strong fit, effectively capturing the relationship between the 95th percentile of posterior inclusion probabilities *PIP_q_*_95_, the coefficient of variation (CV), and sample size. A non-linear regression model was fitted using the Richard Curve^38^, as shown in Figure 1. Parameter estimates were highly significant (p < 0.000001), with a residual standard error of 0.05911. Key parameter values are presented in Table 1.

**Figure 1.**
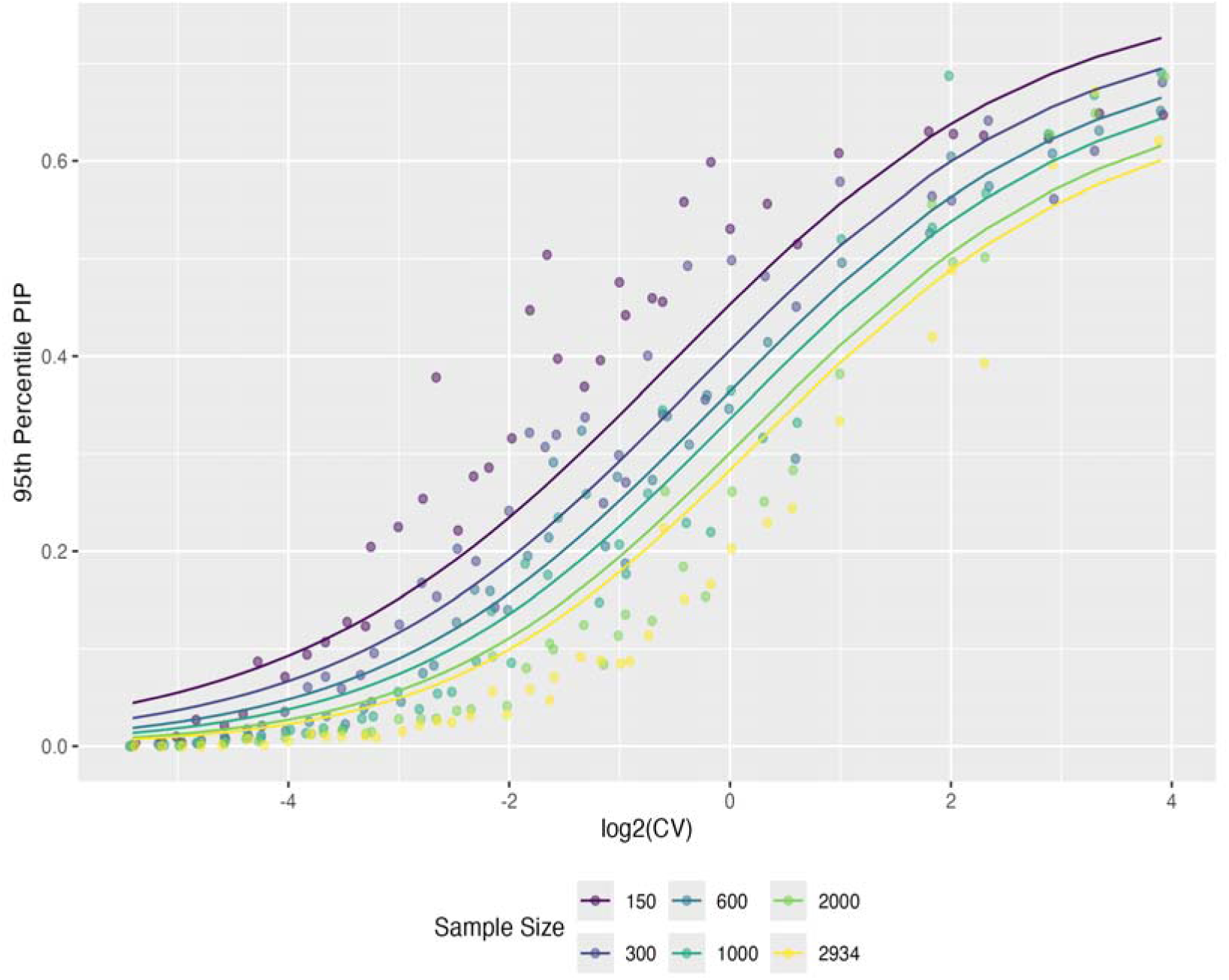
Non-Linear Regression Model Fitted Using the Richard Curve

**Table 1.**
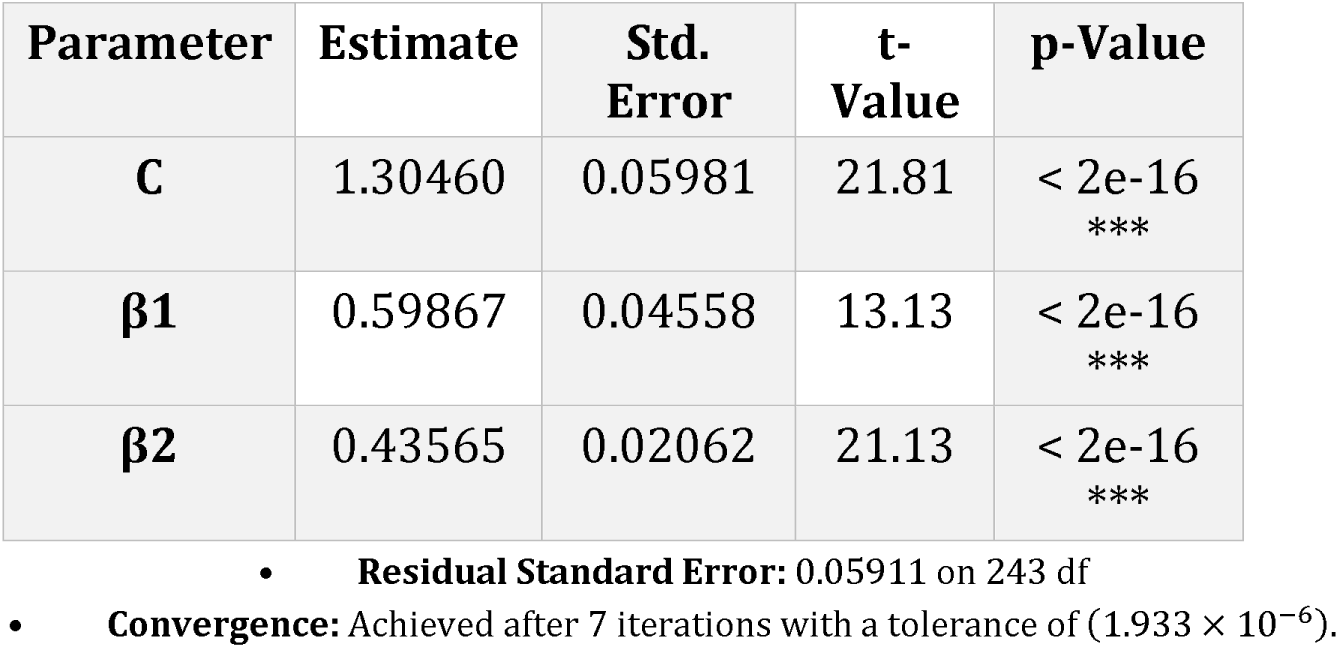
Logistic Model Parameter Estimates

Observed (dots) and predicted *PIP*_*q*95_ values (lines) demonstrated a strong association, shown in Figure 1 that highlighted minimal deviation between the two. The four-parameter logistic model effectively captured nonlinear relationships, but it displayed slight over conservatism for large sample sizes and slight anti-conservatism for smaller sample sizes. These limitations warrant further exploration in future studies.

## Test Size Performance

### New Dynamic Threshold

The dynamically calculated threshold function, based on the logistic model, maintained test sizes closer to the nominal 5% across all conditions. This adaptive approach addressed the inconsistencies of the fixed threshold and provided more reliable statistical inferences. The new threshold function, calculated by fitting a four-parameter logistic model, dynamically adjusted PIP thresholds based on CV and sample size:

Next, we compared the test size for both the fixed (constant at 0.5), and new dynamic thresholds, by evaluating them across all CV and sample size combinations using another 50 simulated datasets (validation datasets).

The graph depicts the relationship between the Coefficient of Variation (CV) (log2-transformed) and test size (y-axis), with a threshold of 0.95 represented by a red dashed line indicating the ideal test size. The graph also incorporates sample size (log10-transformed) as the point size and differentiates between threshold classifications, namely Dynamic and Fixed 0.5, through color and shape. Key observations from the graph include the trend of test sizes clustering near the ideal threshold of 0.95, particularly at lower CV values. As the CV increases, test sizes generally decrease, suggesting a potential loss of statistical power with a higher CV. When comparing the dynamic and fixed thresholding methods, dynamic thresholds (represented by green diamonds) exhibit more variability in test size adjustments, indicating greater adaptability to different conditions. In contrast, fixed thresholds (orange circles) display a more consistent pattern, maintaining a relatively stable distribution of test sizes. The impact of sample size is also evident, as larger sample sizes (larger points) tend to cluster closer to the ideal threshold, supporting the idea that increased sample size helps preserve statistical power. Smaller sample sizes, notably higher CV values, show more deviation from the 0.95 threshold.

The new threshold consistently performed closer to the nominal 5% test size, while the old threshold displayed significant deviations, particularly at extreme CV values or smaller sample sizes.

### Impact of Sample Size and CV

As shown in Figure 2, the test size for both the old and new thresholds was evaluated across all CV and sample size combinations. The fixed PIP threshold of 0.5 exhibited significant variability in test size performance across different CV and sample size combinations. For low CV values, the threshold was overly conservative, resulting in reduced power. In contrast, for high CV values, the threshold was highly anti-conservative, leading to inflated false positive rates. The analysis revealed effects of sample size and CV on *PIP_q_*_95_ :

- Smaller Sample Sizes: Higher *PIP_q_*_95_ values were observed, with greater sensitivity to CV variations.
- Higher CV Values: Test sizes deviated significantly from the nominal 5% under the fixed threshold, underscoring the need for adaptive sensitivity.

**Figure 2.**
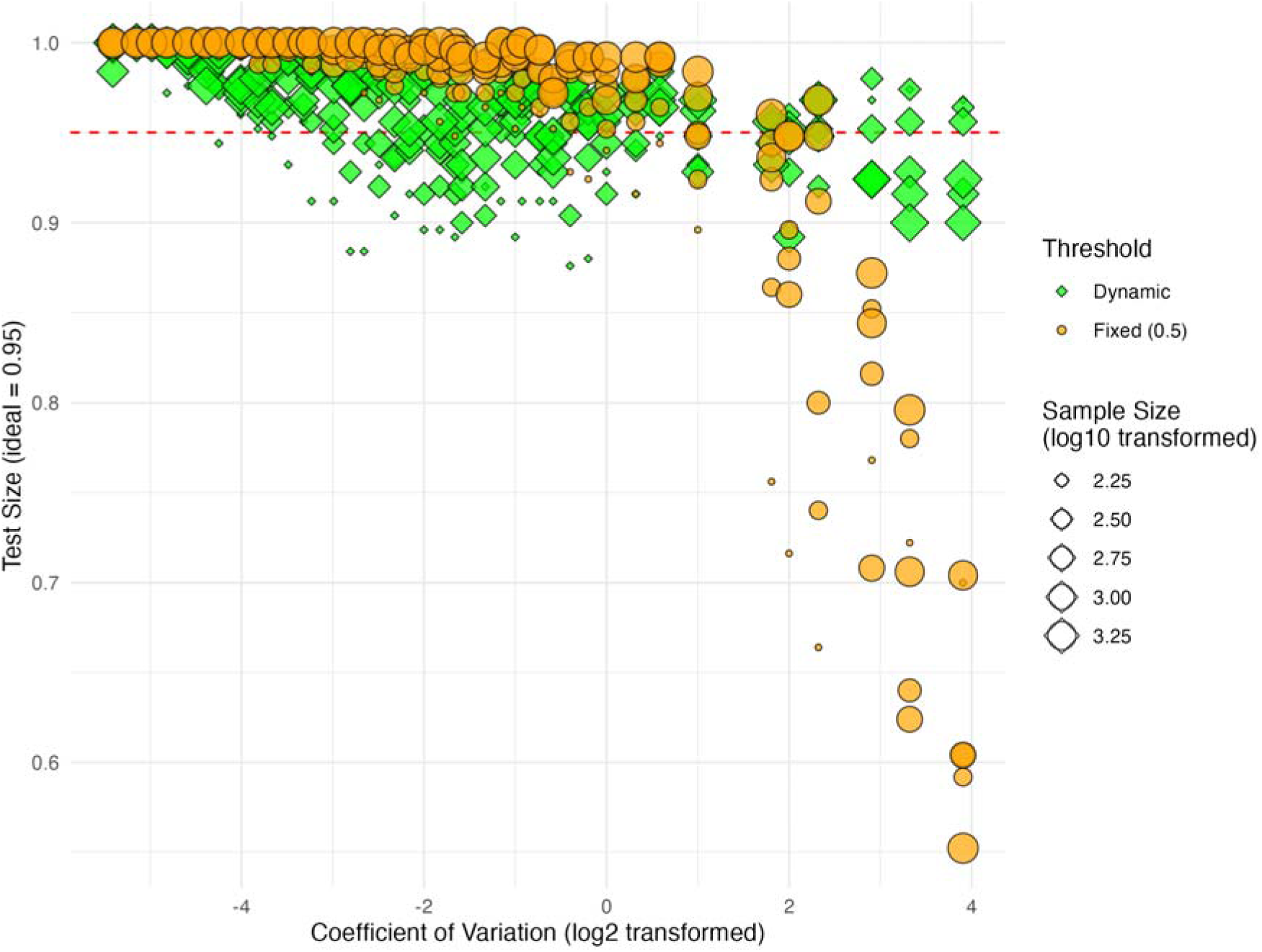
Test Size Comparison Between Fixed and Dynamic Thresholds

The dynamic threshold function effectively compensated for these variations, maintaining consistent performance across diverse conditions. The results showed that dynamic threshold outperformed the fixed threshold (0.5) in test size control and sensitivity. Key findings include:

- Test Size Control: The dynamic threshold consistently achieved test sizes closer to the nominal 5%, regardless of CV and sample size.
- Sensitivity: The dynamic threshold identified a greater number of true associations, particularly for smaller CV values and larger sample sizes.
- Positive Test Identification: The new threshold showed enhanced performance in detecting positive tests without increasing false positives.

The validation data results for various combinations of sample size and CV values are shown in Table 2. Detailed results for other combinations can be found in the supplementary material.

**Table 2:** Sample Validation Data Results for Various Combinations of Sample Size and CV Values

| sampleSize | CV | testSizeOldProp | testSizeNewProp |
| --- | --- | --- | --- |
| 150 | 0.126 | 0.992 | 0.912 |
| 150 | 0.100 | 0.992 | 0.956 |
| 150 | 0.282 | 0.956 | 0.896 |
| 150 | 0.028 | 1.000 | 0.996 |
| 150 | 0.071 | 0.992 | 0.952 |
| 300 | 4.000 | 0.896 | 0.960 |
| **300** | 0.024 | 1.000 | 1.000 |
| **300** | 0.143 | 0.996 | 0.928 |
| **300** | 0.079 | 0.988 | 0.956 |
| **300** | 15.000 | 0.592 | 0.964 |
| **600** | 4.000 | 0.880 | 0.928 |
| **600** | 0.603 | 0.984 | 0.948 |
| **600** | 0.398 | 0.988 | 0.920 |
| **600** | 0.089 | 0.996 | 0.992 |
| **600** | 0.024 | 1.000 | 0.996 |
| **1000** | 7.500 | 0.708 | 0.924 |
| **1000** | 3.500 | 0.944 | 0.944 |
| **1000** | 0.178 | 0.988 | 0.972 |
| **1000** | 0.032 | 1.000 | 1.000 |
| **1000** | 0.667 | 0.972 | 0.928 |
| **2000** | 1.250 | 0.980 | 0.976 |
| **2000** | 4.000 | 0.948 | 0.952 |
| **2000** | 0.126 | 0.996 | 0.976 |
| **2000** | 0.500 | 0.992 | 0.968 |
| **2000** | 0.158 | 1.000 | 0.984 |
| **2934** | 0.500 | 0.996 | 0.968 |
| **2934** | 0.053 | 1.000 | 0.996 |
| **2934** | 0.667 | 0.972 | 0.948 |
| **2934** | 3.500 | 0.960 | 0.956 |
| **2934** | 5.000 | 0.968 | 0.968 |

### Real Data Validation

For the real data analysis and performance demonstration, we used the 2011-2014 National Health and Nutrition Examination Survey (NHANES) data for those 60 or older^44,45^. The initial sample size was 2934, but after omitting the missing values, we ended up with a dataset with a sample size of 949. Separate datasets were acquired: cognitive function assessments and urine metal concentrations.

Cognitive scores (CSCORE) served as the response variable. A cognitive evaluation is a set of tests used to evaluate an individual’s memory, attention, and processing speed. Participants in the NHANES survey who were at least 60 years old, could read and understand one of the stated languages, and did not need a proxy informant to complete the questions, were given this questionnaire. The system consisted of word learning and recall modules from Consortium to Establish a Registry for Alzheimer’s Disease, the Animal Fluency Test, and the Digit Symbol Substitution Test.(citation) Participants who did not answer the cognitive examination were excluded from the results analysis. The education-dependent cognitive z-score is a measure of cognitive performance that takes education level into account. It is calculated by stratifying individuals into three educational levels: less than high school, high school graduate, and greater than high school degree. The education-dependent scores for each cognitive test are then centered and scaled to have a mean of 0 and a standard deviation of 1 within each education stratum. A global cognitive measure is calculated as the average of standardized scores from each cognitive test. This allows for a more accurate assessment of cognitive performance, as education level can significantly impact an individual’s cognitive abilities^46–48^.

Key metals included cadmium, manganese, lead, strontium, tin, uranium, and cobalt available in the NHANES data. All exposure variables were log-transformed and standardized to account for variability and skewness. We then fit the BKMR model to assess the joint effect of metal exposures on the cognitive score. The posterior inclusion probability (PIP) derived from the BKMR model is summarized in Table 3.

**Table 3:**
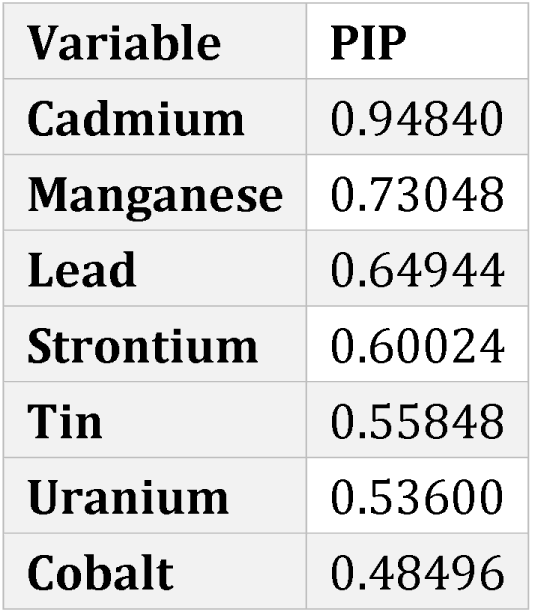
Posterior Inclusion probability (PIP)

We then use the above formula (iii) to determine the cutoff point for the PIP to select the most important metals related to the cognitive score. Here, the response variable CV was 15.18098, and the sample size was 949. Using the formula, we calculated that the appropriate threshold for PIP was 0.6463245. In our analysis, urine cadmium was the most important variable, with a PIP of 0.9484, followed by manganese and lead. If we had used the traditional cutoff value of 0.5 instead of this dynamic threshold, strontium, tin, and uranium would also have been selected as important variables, and only urine cobalt (with a PIP value of 0.48496) would not have been selected. Validation with real datasets displays the new threshold’s superiority in identifying potentially impactful associations.

After BKMR analysis, post-hoc univariate predictor-response relationships were assessed to evaluate the individual effects of each metal on cognitive scores. The results were visualized in Figure 3 as predictor-response functions:

- General Patterns: Exposure to specific metals, such as cadmium and manganese, exhibited nonlinear relationships with cognitive scores.
- Confidence Intervals: The 95% confidence intervals provided insights into the uncertainty around the estimated effects, with narrower intervals observed for more influential metals.
- Variation Across Metals: Some metals had more pronounced effects, while others displayed flat or negligible relationships with the cognitive outcomes.

**Figure 3:**
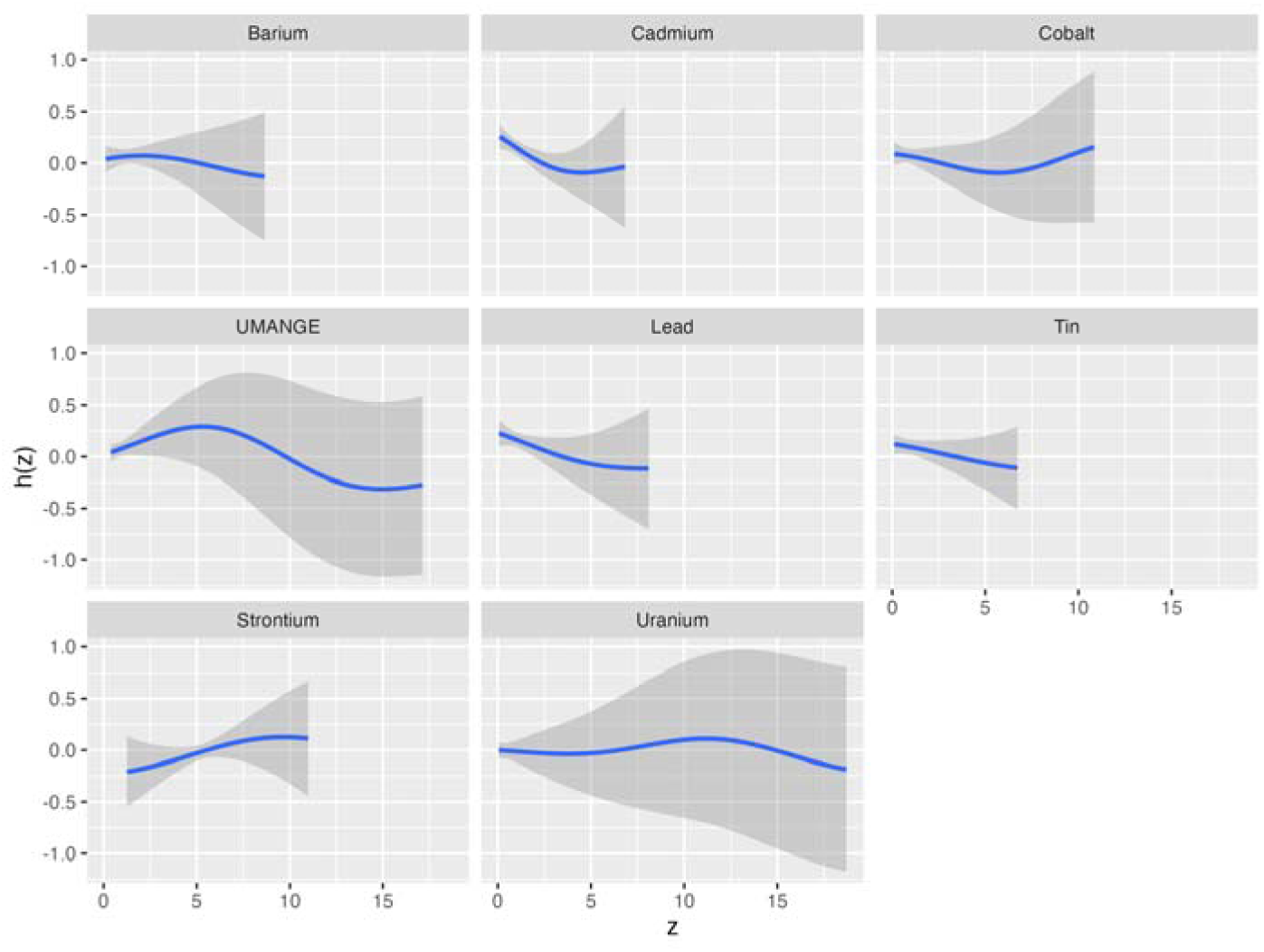
Univariate predictor-response functions showing the estimated effects (h(z)) of each metal on cognitive scores, with 95% confidence intervals.

Univariate predictor-response relationships revealed nonlinear associations between individual metals and cognitive scores. Metals such as cadmium and manganese exhibited pronounced effects with narrower confidence intervals, reflecting robust and consistent associations with cognitive outcomes. Other metals, such as uranium, displayed negligible effects, underscoring the variability in metal-specific impacts on cognitive performance. Cadmium, manganese, and lead are toxic heavy metals that have been linked to adverse effects on cognitive function^49^. Exposure to the cadmium has been associated with impaired neurodevelopment in children and cognitive decline in adults^50^. Manganese, when accumulated in the brain, can lead to neurotoxicity, particularly affecting the basal ganglia, and has been linked to cognitive deficits, including memory and attention impairments^51^. Lead exposure, especially in early childhood, is known to cause a range of cognitive deficits, including lower IQ, attention problems, and learning disabilities^52^. These metals can interfere with normal neural function through oxidative stress, inflammation, and disruption of synaptic plasticity^49–54^.

Overall risk summaries in Figure 4 quantified the combined effect of all urinary metals on cognitive performance. The analysis focused on changes in cognitive scores across quantiles of metal mixture exposure:

- Quantile-Based Risk Analysis: Cognitive scores were assessed at various quantiles of exposure (25th to 75th percentile) using the median exposure level as a reference.
- Findings: Increased overall metal exposure was associated with a significant reduction in cognitive scores, with the effect being most pronounced at higher exposure quantiles.
- Confidence Intervals: Point estimates of risk were presented alongside 95% confidence intervals, providing a clear visualization of the exposure-response relationship.

**Figure 4:**
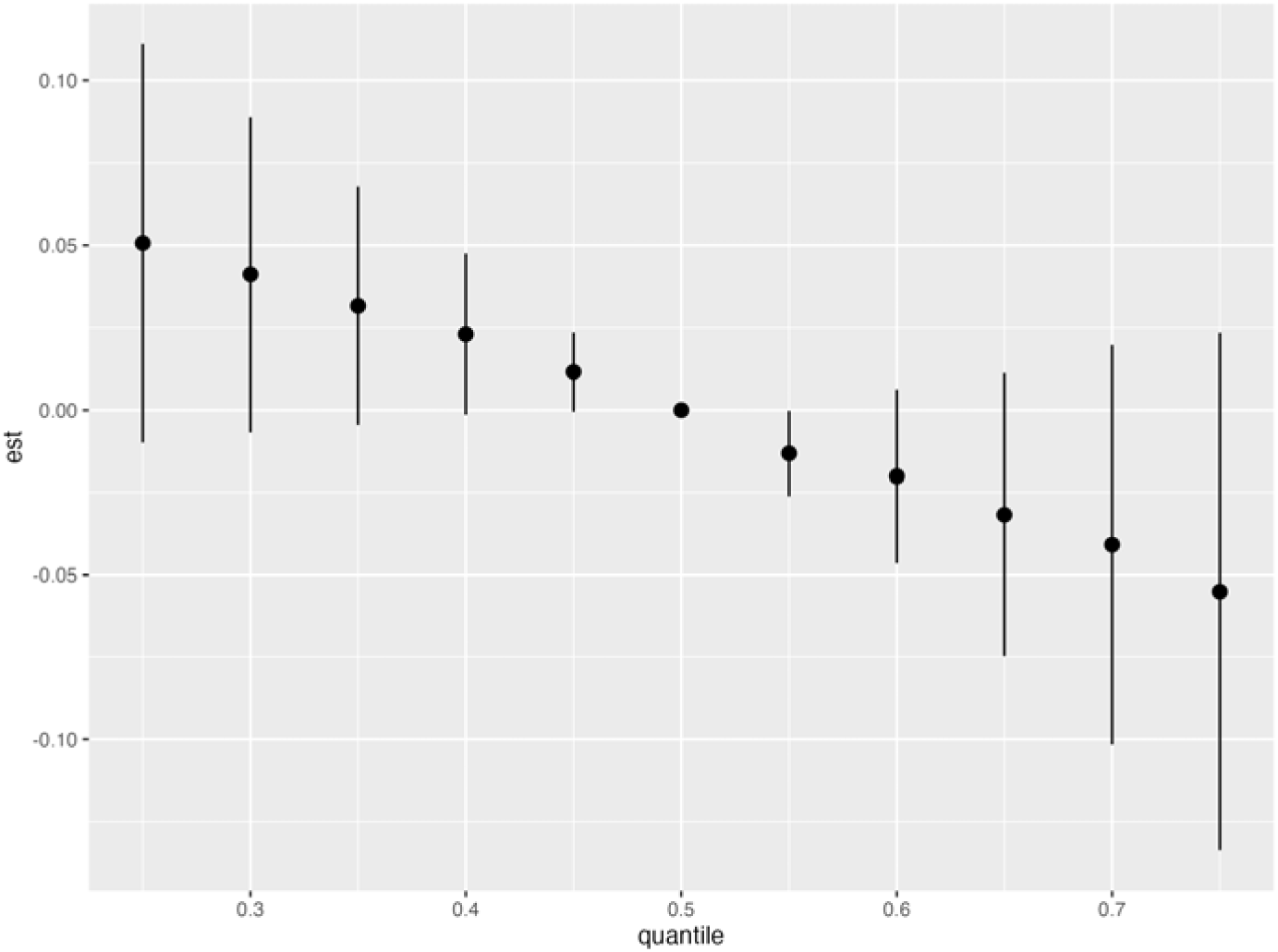
Overall risk summaries displaying estimated cognitive score changes across exposure quantiles, with error bars representing 95% confidence intervals.

These findings highlight the strength of BKMR in capturing complex and nonlinear exposure-response relationships, which are often challenging to model with traditional linear regression techniques. Further, these results highlight the utility of the BKMR updated threshold in disentangling the complex interactions between environmental exposures and health outcomes, providing valuable insights for environmental health research and public health policy. Validation with real-world datasets reinforced the superiority of the dynamic threshold function over the traditional fixed threshold. The dynamic threshold consistently identified true associations while maintaining appropriate test size control, confirming its practical applicability. These findings highlight the necessity of adaptive thresholds for improving the reliability of statistical tests in high-dimensional and variable-rich datasets.

## Discussion

This study presents a significant advancement in applying Bayesian Kernel Machine Regression (BKMR) by introducing a dynamic threshold function for posterior inclusion probability (PIP). Unlike the conventional fixed-threshold approach, which assumes a static PIP threshold (typically 0.5), our dynamic threshold adapts to key dataset characteristics, including the coefficient of variation (CV) and sample size. This adaptability addresses critical limitations in conventional BKMR applications, particularly in high-dimensional and variable-rich datasets, such as those encountered in environmental health research, where exposure-response relationships are often nonlinear and influenced by complex interdependencies.

Our results demonstrate that the conventional fixed PIP threshold yields substantial variability in test size performance. Specifically, our simulations reveal that fixed-threshold BKMR is overly conservative for low CV values, reducing statistical power and potentially overlooking important variables. Conversely, for high CV values, the fixed threshold becomes overly liberal, leading to inflated false-positive rates. This variability poses challenges in identifying genuine exposure-response associations, especially in datasets with small sample sizes or high response variability. By contrast, modelled using a four-parameter logistic function, our dynamic threshold function adjusts PIP thresholds based on dataset-specific parameters, maintaining test sizes close to the nominal 5% across various conditions. This ensures a more stable and reliable selection of variables, reducing Type I and Type II errors. The effectiveness of the dynamic threshold function was validated through extensive simulations, where it consistently outperformed the fixed threshold in test size control and sensitivity. The dynamic threshold was particularly beneficial for extreme CV values and smaller sample sizes—scenarios where the fixed threshold exhibited the most substantial deviations. These findings reinforce the necessity of an adaptive thresholding approach in high-dimensional epidemiological studies. Our findings align with and extend prior research on BKMR and variable selection in high-dimensional modelling. Studies such as those by Bobb et al. (2015) and Zigler and Dominici (2014) demonstrated BKMR’s effectiveness in capturing nonlinear exposure-response relationships^5,6,8,12–14^. However, they relied on a fixed PIP threshold for variable selection. More recent studies (Wilson et al., 2018; Valeri et al., 2017) have challenged this fixed-threshold assumption, citing its inability to account for dataset-specific variability[[^55^;^11^;^56^]. Our study empirically confirms these concerns, showing that static thresholds lead to inconsistent test size performance.

Alternative variable selection strategies have been explored in the literature. Liu et al. introduced weighting schemes for PIP calculation to enhance sensitivity in high-dimensional models^57^, while other researcher proposed hierarchical Bayesian frameworks to adjust for response variability^58,59^. Although these methods improved statistical performance relative to fixed-threshold BKMR, they lacked an explicit mechanism for adapting to sample size and CV variations. Our logistic model-based dynamic threshold approach builds upon this work by offering a transparent, interpretable, and robust framework that ensures reliable test size control across diverse conditions.

To assess the practical applicability of our approach, we applied the dynamic threshold to real-world data from the National Health and Nutrition Examination Survey (NHANES) (2011–2014), focusing on urinary metal exposures and cognitive outcomes in older adults. The results highlighted cadmium, manganese, and lead as significant contributors to cognitive decline—findings well supported by prior epidemiological research^49,60^. Notably, the dynamic threshold excluded strontium and tin, which lack strong evidence for neurotoxicity, whereas the fixed threshold would have incorrectly included them as significant variables. This underscores a key advantage of the dynamic approach: reducing false positives while preserving the integrity of variable selection. By refining exposure-response modeling, our method minimizes the risk of spurious associations and enhances the reliability of findings in environmental health research.

Furthermore, our cumulative risk analyses revealed a dose-response relationship between urinary metal exposures and cognitive decline, particularly in higher exposure quantiles. These findings underscore the broader implications of mental exposures for cognitive health in ageing populations and highlight the need for targeted public health interventions to mitigate these risks.

The dynamic threshold function represents a significant step forward in BKMR applications, providing a more reliable and adaptive framework for variable selection. However, some limitations warrant further investigation. Our simulations suggest that the logistic model is slightly conservative for large sample sizes and slightly liberal for smaller sample sizes, leading to minor biases in threshold estimation under extreme conditions. Future research should refine the dynamic threshold formula to ensure optimal performance across all sample sizes and dataset conditions. Additionally, our study was limited to a fixed number of simulated metal exposures, preventing us from assessing the impact of an asymptotically increasing dimensionality. As computational resources become more powerful and accessible, future work should explore how response CV, sample size, and predictor dimensionality influence PIP thresholds in high-dimensional models. Expanding our approach to incorporate additional linear covariates, such as demographic and socioeconomic factors, could also improve precision and contextual relevance in exposure-response analyses.

Building on this study’s strengths, future research should focus on increasing the variety of simulation studies and extending the dynamic threshold approach to other datasets and domains. Beyond environmental health research, the dynamic threshold framework has potential applications in other high-dimensional fields, including genomics, metabolomics, and air pollution epidemiology. Extending this work to longitudinal studies would allow for the exploration of cumulative and long-term exposure effects, providing deeper insights into complex mixtures’ health impacts over time.

## Conclusion

This study demonstrates the effectiveness of a dynamic threshold approach in improving the reliability and sensitivity of BKMR analyses. By accounting for key dataset characteristics such as CV and sample size, the dynamic threshold function addresses the limitations of fixed thresholds, enabling more accurate identification of significant exposure-response relationships. These advancements significantly impact environmental health research, providing a robust methodological foundation for analyzing complex, high-dimensional datasets. The dynamic threshold enhances the precision of variable selection. It underscores the potential of adaptive methods in addressing the challenges of modern data analysis, ultimately contributing to more informed public health interventions and policies.

## AUTHOR CONTRIBUTIONS

All co-authors have meaningfully contributed to the production of this manuscript including funding, conception, data analysis, edition, revision, and final approval of the manuscript.

## CONFLICT OF INTEREST

The authors declare no potential conflict of interests. Data and Code Availability

The code and simulation data will be made available upon acceptance.

## Supplementary Material

Supplementary martial is attached as a separate file

## Data Availability

The code and simulation data will be made available upon reasonable request to the authors.

