## Supplemental File for "Enhancing Bayesian Kernel Machine Regression: A Dynamic Thresholding Framework to Address Variability and Skewness in High-Dimensional Environmental Health Data"

| sampleSize | CV | testSizeOld | testSizeNew | testSizeOldProp | testSizeNewProp |
| --- | --- | --- | --- | --- | --- |
| 150 | 0.0235294117647059 | 250 | 249 | 1 | 0.996 |
| 150 | 0.02818383 | 250 | 249 | 1 | 0.996 |
| 150 | 0.03162278 | 250 | 250 | 1 | 1 |
| 150 | 0.03548134 | 250 | 243 | 1 | 0.972 |
| 150 | 0.0416666666666667 | 250 | 244 | 1 | 0.976 |
| 150 | 0.04786301 | 249 | 245 | 0.996 | 0.98 |
| 150 | 0.0526315789473684 | 248 | 236 | 0.992 | 0.944 |
| 150 | 0.062111801242236 | 248 | 240 | 0.992 | 0.96 |
| 150 | 0.07079458 | 248 | 238 | 0.992 | 0.952 |
| 150 | 0.078740157480315 | 248 | 237 | 0.992 | 0.948 |
| 150 | 0.08912509 | 248 | 233 | 0.992 | 0.932 |
| 150 | 0.1 | 248 | 239 | 0.992 | 0.956 |
| 150 | 0.106382978723404 | 246 | 228 | 0.984 | 0.912 |
| 150 | 0.1258925 | 248 | 228 | 0.992 | 0.912 |
| 150 | 0.142857142857143 | 249 | 221 | 0.996 | 0.884 |
| 150 | 0.1584893 | 243 | 221 | 0.972 | 0.884 |
| 150 | 0.1778279 | 242 | 233 | 0.968 | 0.932 |
| 150 | 0.2 | 246 | 226 | 0.984 | 0.904 |
| 150 | 0.2238721 | 246 | 229 | 0.984 | 0.916 |
| 150 | 0.25 | 243 | 224 | 0.972 | 0.896 |
| 150 | 0.2818383 | 239 | 224 | 0.956 | 0.896 |
| 150 | 0.3162278 | 237 | 223 | 0.948 | 0.892 |
| 150 | 0.333333333333333 | 243 | 231 | 0.972 | 0.924 |
| 150 | 0.3981072 | 241 | 230 | 0.964 | 0.92 |
| 150 | 0.448430493273543 | 243 | 228 | 0.972 | 0.912 |
| 150 | 0.5 | 238 | 223 | 0.952 | 0.892 |
| 150 | 0.526315789473684 | 241 | 228 | 0.964 | 0.912 |
| 150 | 0.6025596 | 240 | 228 | 0.96 | 0.912 |
| 150 | 0.666666666666667 | 238 | 228 | 0.952 | 0.912 |
| 150 | 0.7585776 | 232 | 219 | 0.928 | 0.876 |
| 150 | 0.8709636 | 231 | 220 | 0.924 | 0.88 |
| 150 | 1 | 235 | 232 | 0.94 | 0.928 |
| 150 | 1.25 | 229 | 229 | 0.916 | 0.916 |
| 150 | 1.5 | 236 | 237 | 0.944 | 0.948 |
| 150 | 2 | 448 | 465 | 0.896 | 0.93 |
| 150 | 3.5 | 189 | 235 | 0.756 | 0.94 |
| 150 | 4 | 179 | 238 | 0.716 | 0.952 |
| 150 | 5 | 166 | 239 | 0.664 | 0.956 |
| 150 | 7.5 | 192 | 242 | 0.768 | 0.968 |
| 150 | 10 | 361 | 487 | 0.722 | 0.974 |
| 150 | 15 | 175 | 242 | 0.7 | 0.968 |
| 300 | 0.0235294117647059 | 250 | 250 | 1 | 1 |
| 300 | 0.02818383 | 250 | 250 | 1 | 1 |
| 300 | 0.03162278 | 250 | 250 | 1 | 1 |
| 300 | 0.03548134 | 250 | 249 | 1 | 0.996 |
| 300 | 0.0416666666666667 | 250 | 247 | 1 | 0.988 |
| 300 | 0.04786301 | 250 | 246 | 1 | 0.984 |
| 300 | 0.0526315789473684 | 250 | 244 | 1 | 0.976 |
| 300 | 0.062111801242236 | 249 | 243 | 0.996 | 0.972 |
| 300 | 0.07079458 | 247 | 240 | 0.988 | 0.96 |
| 300 | 0.078740157480315 | 247 | 239 | 0.988 | 0.956 |
| 300 | 0.08912509 | 250 | 241 | 1 | 0.964 |
| 300 | 0.1 | 250 | 246 | 1 | 0.984 |
| 300 | 0.106382978723404 | 247 | 239 | 0.988 | 0.956 |
| 300 | 0.1258925 | 248 | 236 | 0.992 | 0.944 |
| 300 | 0.142857142857143 | 249 | 232 | 0.996 | 0.928 |
| 300 | 0.1584893 | 248 | 236 | 0.992 | 0.944 |
| 300 | 0.1778279 | 246 | 230 | 0.984 | 0.92 |
| 300 | 0.2 | 244 | 234 | 0.976 | 0.936 |
| 300 | 0.2238721 | 249 | 242 | 0.996 | 0.968 |
| 300 | 0.25 | 246 | 229 | 0.984 | 0.916 |
| 300 | 0.2818383 | 245 | 230 | 0.98 | 0.92 |
| 300 | 0.3162278 | 243 | 229 | 0.972 | 0.916 |
| 300 | 0.333333333333333 | 243 | 225 | 0.972 | 0.9 |
| 300 | 0.3981072 | 243 | 226 | 0.972 | 0.904 |
| 300 | 0.448430493273543 | 249 | 241 | 0.996 | 0.964 |
| 300 | 0.5 | 243 | 237 | 0.972 | 0.948 |
| 300 | 0.526315789473684 | 245 | 240 | 0.98 | 0.96 |
| 300 | 0.6025596 | 241 | 233 | 0.964 | 0.932 |
| 300 | 0.666666666666667 | 243 | 237 | 0.972 | 0.948 |
| 300 | 0.7585776 | 239 | 226 | 0.956 | 0.904 |
| 300 | 0.8709636 | 241 | 238 | 0.964 | 0.952 |
| 300 | 1 | 238 | 229 | 0.952 | 0.916 |
| 300 | 1.25 | 239 | 235 | 0.956 | 0.94 |
| 300 | 1.5 | 241 | 240 | 0.964 | 0.96 |
| 300 | 2 | 462 | 466 | 0.924 | 0.932 |
| 300 | 3.5 | 216 | 239 | 0.864 | 0.956 |
| 300 | 4 | 224 | 240 | 0.896 | 0.96 |
| 300 | 5 | 185 | 230 | 0.74 | 0.92 |
| 300 | 7.5 | 213 | 245 | 0.852 | 0.98 |
| 300 | 10 | 390 | 487 | 0.78 | 0.974 |
| 300 | 15 | 148 | 241 | 0.592 | 0.964 |
| 600 | 0.0235294117647059 | 250 | 249 | 1 | 0.996 |
| 600 | 0.02818383 | 250 | 250 | 1 | 1 |
| 600 | 0.03162278 | 250 | 250 | 1 | 1 |
| 600 | 0.03548134 | 250 | 250 | 1 | 1 |
| 600 | 0.0416666666666667 | 250 | 248 | 1 | 0.992 |
| 600 | 0.04786301 | 250 | 248 | 1 | 0.992 |
| 600 | 0.0526315789473684 | 250 | 244 | 1 | 0.976 |
| 600 | 0.062111801242236 | 250 | 245 | 1 | 0.98 |
| 600 | 0.07079458 | 250 | 243 | 1 | 0.972 |
| 600 | 0.078740157480315 | 250 | 242 | 1 | 0.968 |
| 600 | 0.08912509 | 249 | 248 | 0.996 | 0.992 |
| 600 | 0.1 | 250 | 241 | 1 | 0.964 |
| 600 | 0.106382978723404 | 250 | 245 | 1 | 0.98 |
| 600 | 0.1258925 | 249 | 238 | 0.996 | 0.952 |
| 600 | 0.142857142857143 | 249 | 243 | 0.996 | 0.972 |
| 600 | 0.1584893 | 248 | 241 | 0.992 | 0.964 |
| 600 | 0.1778279 | 249 | 236 | 0.996 | 0.944 |
| 600 | 0.2 | 246 | 234 | 0.984 | 0.936 |
| 600 | 0.2238721 | 248 | 235 | 0.992 | 0.94 |
| 600 | 0.25 | 249 | 238 | 0.996 | 0.952 |
| 600 | 0.2818383 | 248 | 235 | 0.992 | 0.94 |
| 600 | 0.3162278 | 248 | 233 | 0.992 | 0.932 |
| 600 | 0.333333333333333 | 247 | 230 | 0.988 | 0.92 |
| 600 | 0.3981072 | 247 | 230 | 0.988 | 0.92 |
| 600 | 0.448430493273543 | 246 | 238 | 0.984 | 0.952 |
| 600 | 0.5 | 248 | 233 | 0.992 | 0.932 |
| 600 | 0.526315789473684 | 248 | 245 | 0.992 | 0.98 |
| 600 | 0.6025596 | 246 | 237 | 0.984 | 0.948 |
| 600 | 0.666666666666667 | 245 | 236 | 0.98 | 0.944 |
| 600 | 0.7585776 | 246 | 239 | 0.984 | 0.956 |
| 600 | 0.8709636 | 247 | 234 | 0.988 | 0.936 |
| 600 | 1 | 244 | 239 | 0.976 | 0.956 |
| 600 | 1.25 | 242 | 236 | 0.968 | 0.944 |
| 600 | 1.5 | 247 | 246 | 0.988 | 0.984 |
| 600 | 2 | 475 | 474 | 0.95 | 0.948 |
| 600 | 3.5 | 231 | 239 | 0.924 | 0.956 |
| 600 | 4 | 220 | 232 | 0.88 | 0.928 |
| 600 | 5 | 200 | 238 | 0.8 | 0.952 |
| 600 | 7.5 | 204 | 238 | 0.816 | 0.952 |
| 600 | 10 | 320 | 478 | 0.64 | 0.956 |
| 600 | 15 | 151 | 239 | 0.604 | 0.956 |
| 1000 | 0.0235294117647059 | 250 | 246 | 1 | 0.984 |
| 1000 | 0.02818383 | 250 | 250 | 1 | 1 |
| 1000 | 0.03162278 | 250 | 250 | 1 | 1 |
| 1000 | 0.03548134 | 250 | 250 | 1 | 1 |
| 1000 | 0.0416666666666667 | 250 | 247 | 1 | 0.988 |
| 1000 | 0.04786301 | 250 | 247 | 1 | 0.988 |
| 1000 | 0.0526315789473684 | 250 | 248 | 1 | 0.992 |
| 1000 | 0.062111801242236 | 250 | 242 | 1 | 0.968 |
| 1000 | 0.07079458 | 250 | 246 | 1 | 0.984 |
| 1000 | 0.078740157480315 | 250 | 246 | 1 | 0.984 |
| 1000 | 0.08912509 | 250 | 245 | 1 | 0.98 |
| 1000 | 0.1 | 250 | 242 | 1 | 0.968 |
| 1000 | 0.106382978723404 | 250 | 245 | 1 | 0.98 |
| 1000 | 0.1258925 | 247 | 240 | 0.988 | 0.96 |
| 1000 | 0.142857142857143 | 250 | 244 | 1 | 0.976 |
| 1000 | 0.1584893 | 249 | 245 | 0.996 | 0.98 |
| 1000 | 0.1778279 | 247 | 243 | 0.988 | 0.972 |
| 1000 | 0.2 | 248 | 239 | 0.992 | 0.956 |
| 1000 | 0.2238721 | 249 | 236 | 0.996 | 0.944 |
| 1000 | 0.25 | 250 | 245 | 1 | 0.98 |
| 1000 | 0.2818383 | 246 | 236 | 0.984 | 0.944 |
| 1000 | 0.3162278 | 247 | 236 | 0.988 | 0.944 |
| 1000 | 0.333333333333333 | 246 | 233 | 0.984 | 0.932 |
| 1000 | 0.3981072 | 246 | 234 | 0.984 | 0.936 |
| 1000 | 0.448430493273543 | 247 | 242 | 0.988 | 0.968 |
| 1000 | 0.5 | 250 | 239 | 1 | 0.956 |
| 1000 | 0.526315789473684 | 248 | 244 | 0.992 | 0.976 |
| 1000 | 0.6025596 | 246 | 237 | 0.984 | 0.948 |
| 1000 | 0.666666666666667 | 243 | 232 | 0.972 | 0.928 |
| 1000 | 0.7585776 | 248 | 242 | 0.992 | 0.968 |
| 1000 | 0.8709636 | 246 | 241 | 0.984 | 0.964 |
| 1000 | 1 | 246 | 236 | 0.984 | 0.944 |
| 1000 | 1.25 | 245 | 242 | 0.98 | 0.968 |
| 1000 | 1.5 | 247 | 242 | 0.988 | 0.968 |
| 1000 | 2 | 474 | 464 | 0.948 | 0.928 |
| 1000 | 3.5 | 236 | 236 | 0.944 | 0.944 |
| 1000 | 4 | 215 | 223 | 0.86 | 0.892 |
| 1000 | 5 | 228 | 237 | 0.912 | 0.948 |
| 1000 | 7.5 | 177 | 231 | 0.708 | 0.924 |
| 1000 | 10 | 312 | 464 | 0.624 | 0.928 |
| 1000 | 15 | 151 | 229 | 0.604 | 0.916 |
| 2000 | 0.0235294117647059 | 250 | 250 | 1 | 1 |
| 2000 | 0.02818383 | 250 | 250 | 1 | 1 |
| 2000 | 0.03162278 | 250 | 250 | 1 | 1 |
| 2000 | 0.03548134 | 250 | 249 | 1 | 0.996 |
| 2000 | 0.0416666666666667 | 250 | 248 | 1 | 0.992 |
| 2000 | 0.04786301 | 250 | 246 | 1 | 0.984 |
| 2000 | 0.0526315789473684 | 250 | 248 | 1 | 0.992 |
| 2000 | 0.062111801242236 | 250 | 243 | 1 | 0.972 |
| 2000 | 0.07079458 | 250 | 242 | 1 | 0.968 |
| 2000 | 0.078740157480315 | 250 | 241 | 1 | 0.964 |
| 2000 | 0.08912509 | 250 | 248 | 1 | 0.992 |
| 2000 | 0.1 | 250 | 248 | 1 | 0.992 |
| 2000 | 0.106382978723404 | 250 | 246 | 1 | 0.984 |
| 2000 | 0.1258925 | 249 | 244 | 0.996 | 0.976 |
| 2000 | 0.142857142857143 | 248 | 244 | 0.992 | 0.976 |
| 2000 | 0.1584893 | 250 | 246 | 1 | 0.984 |
| 2000 | 0.1778279 | 250 | 248 | 1 | 0.992 |
| 2000 | 0.2 | 250 | 247 | 1 | 0.988 |
| 2000 | 0.2238721 | 249 | 238 | 0.996 | 0.952 |
| 2000 | 0.25 | 250 | 247 | 1 | 0.988 |
| 2000 | 0.2818383 | 248 | 243 | 0.992 | 0.972 |
| 2000 | 0.3162278 | 250 | 238 | 1 | 0.952 |
| 2000 | 0.333333333333333 | 249 | 240 | 0.996 | 0.96 |
| 2000 | 0.3981072 | 247 | 238 | 0.988 | 0.952 |
| 2000 | 0.448430493273543 | 250 | 249 | 1 | 0.996 |
| 2000 | 0.5 | 248 | 242 | 0.992 | 0.968 |
| 2000 | 0.526315789473684 | 250 | 242 | 1 | 0.968 |
| 2000 | 0.6025596 | 249 | 245 | 0.996 | 0.98 |
| 2000 | 0.666666666666667 | 245 | 234 | 0.98 | 0.936 |
| 2000 | 0.7585776 | 247 | 244 | 0.988 | 0.976 |
| 2000 | 0.8709636 | 248 | 241 | 0.992 | 0.964 |
| 2000 | 1 | 242 | 240 | 0.968 | 0.96 |
| 2000 | 1.25 | 245 | 244 | 0.98 | 0.976 |
| 2000 | 1.5 | 248 | 243 | 0.992 | 0.972 |
| 2000 | 2 | 485 | 481 | 0.97 | 0.962 |
| 2000 | 3.5 | 234 | 233 | 0.936 | 0.932 |
| 2000 | 4 | 237 | 238 | 0.948 | 0.952 |
| 2000 | 5 | 237 | 242 | 0.948 | 0.968 |
| 2000 | 7.5 | 211 | 231 | 0.844 | 0.924 |
| 2000 | 10 | 353 | 458 | 0.706 | 0.916 |
| 2000 | 15 | 176 | 225 | 0.704 | 0.9 |
| 2934 | 0.0235294117647059 | 250 | 250 | 1 | 1 |
| 2934 | 0.02818383 | 250 | 250 | 1 | 1 |
| 2934 | 0.03162278 | 250 | 250 | 1 | 1 |
| 2934 | 0.03548134 | 250 | 249 | 1 | 0.996 |
| 2934 | 0.0416666666666667 | 250 | 249 | 1 | 0.996 |
| 2934 | 0.04786301 | 250 | 244 | 1 | 0.976 |
| 2934 | 0.0526315789473684 | 250 | 249 | 1 | 0.996 |
| 2934 | 0.062111801242236 | 250 | 244 | 1 | 0.976 |
| 2934 | 0.07079458 | 250 | 245 | 1 | 0.98 |
| 2934 | 0.078740157480315 | 250 | 247 | 1 | 0.988 |
| 2934 | 0.08912509 | 250 | 248 | 1 | 0.992 |
| 2934 | 0.1 | 250 | 249 | 1 | 0.996 |
| 2934 | 0.106382978723404 | 250 | 248 | 1 | 0.992 |
| 2934 | 0.1258925 | 250 | 247 | 1 | 0.988 |
| 2934 | 0.142857142857143 | 250 | 244 | 1 | 0.976 |
| 2934 | 0.1584893 | 250 | 247 | 1 | 0.988 |
| 2934 | 0.1778279 | 249 | 245 | 0.996 | 0.98 |
| 2934 | 0.2 | 249 | 244 | 0.996 | 0.976 |
| 2934 | 0.2238721 | 248 | 244 | 0.992 | 0.976 |
| 2934 | 0.25 | 249 | 246 | 0.996 | 0.984 |
| 2934 | 0.2818383 | 250 | 239 | 1 | 0.956 |
| 2934 | 0.3162278 | 249 | 246 | 0.996 | 0.984 |
| 2934 | 0.333333333333333 | 248 | 242 | 0.992 | 0.968 |
| 2934 | 0.3981072 | 248 | 241 | 0.992 | 0.964 |
| 2934 | 0.448430493273543 | 250 | 244 | 1 | 0.976 |
| 2934 | 0.5 | 249 | 242 | 0.996 | 0.968 |
| 2934 | 0.526315789473684 | 250 | 244 | 1 | 0.976 |
| 2934 | 0.6025596 | 249 | 243 | 0.996 | 0.972 |
| 2934 | 0.666666666666667 | 243 | 237 | 0.972 | 0.948 |
| 2934 | 0.7585776 | 248 | 244 | 0.992 | 0.976 |
| 2934 | 0.8709636 | 248 | 239 | 0.992 | 0.956 |
| 2934 | 1 | 248 | 243 | 0.992 | 0.972 |
| 2934 | 1.25 | 248 | 241 | 0.992 | 0.964 |
| 2934 | 1.5 | 248 | 241 | 0.992 | 0.964 |
| 2934 | 2 | 492 | 484 | 0.984 | 0.968 |
| 2934 | 3.5 | 240 | 239 | 0.96 | 0.956 |
| 2934 | 4 | 237 | 237 | 0.948 | 0.948 |
| 2934 | 5 | 242 | 242 | 0.968 | 0.968 |
| 2934 | 7.5 | 218 | 231 | 0.872 | 0.924 |
| 2934 | 10 | 398 | 450 | 0.796 | 0.9 |
| 2934 | 15 | 138 | 231 | 0.552 | 0.924 |
